# Spatial and Temporal Evaluation of Violent and Total Crime Following Community-Led Urban Greening in Indianapolis

**DOI:** 10.64898/2026.10.19.26357148

**Authors:** Jiyeon Song, Yi Li

## Abstract

Community-led urban greening may contribute to neighborhood safety, but its relationship with crime remains uncertain. We examined whether Keep Indianapolis Beautiful Project GreenSpaces in Indianapolis were associated with changes in violence and total crime. We used a matched-pair difference-in-differences design comparing funded sites with matched unfunded applicant sites. The primary analysis included 29 matched pairs with two years of pre- and post-grant follow-up. Monthly Part I violent offenses and total crime within 600-m buffers were modeled using Poisson pseudo-maximum likelihood. In the first post-grant year, the violent-crime estimate was lower but imprecise (IRR, 0.903; 95% CI, 0.805-1.014). In the second post-grant year, the relative rate was lower (IRR, 0.830; 95% CI, 0.718-0.961). Total crime estimates showed no clear post-grant change. Additional analyses indicated some sensitivity to spatial scale, but no sustained divergence in pre-grant crime patterns. Analyses of short-term crime dependence showed clear month-to-month persistence in violent crime within sites but limited evidence of stable spatial dependence across nearby sites. Therefore, within the two-year post-grant period examined, Project GreenSpaces was associated most consistently with lower relative violent-crime rates in the second year, while no corresponding decrease was observed for total recorded crime. Longer follow-up is needed to determine whether these patterns persist or change as projects mature.

## 1. Introduction

Criminological research has long focused on understanding the complexities of crime, including its patterns, causes, and potential intervention approaches [1, 2]. More recently, the intersection of crime research and urban greening has received increasing attention as cities seek place-based approaches to improve neighborhood environments and community well-being [3–5]. A growing body of literature suggests that greening vacant or underutilized urban spaces may be associated with reductions in crime and violence, although findings have varied across settings, intervention types, and study designs [6–8]. These potential crime-reducing effects may operate through both physical remediation which can mitigate cues of disorder and improve sightlines, and social activation that encourages community use [9–11].

However, the complex interaction of neighborhood and environmental factors frequently hinders the identification of conclusive results in these studies. Existing studies have faced challenges in fully accounting for confounders such as socioeconomic status and demographic characteristics that often differ systematically between green neighborhoods and control areas [12–14]. This concern is particularly relevant when greening projects are not randomly assigned: neighborhoods that receive or pursue environmental improvements may already possess unobserved community characteristics that independently influence crime [15, 16]. Comparisons of crime levels observed after greening may therefore confound pre-existing neighborhood differences with changes associated with the intervention.

In this study, we utilized a dataset of community-led greening interventions implemented by Keep Indianapolis Beautiful (KIB), a nonprofit organization dedicated to aesthetic and ecological improvement in Indianapolis, Indiana. Specifically, we examined Project GreenSpaces, an initiative that transforms vacant lots into functional community spaces [17]. Unlike a uniform municipal landscaping intervention, the KIB model relies on community participation.

Neighborhood groups apply for the program, participate in designing the space, and contribute to its ongoing maintenance and care. Thus, Project GreenSpaces represents a community-engaged greening program that combines physical site improvement with continued community involvement.

To assess whether these community-driven projects were followed by changes in urban safety, the primary objective of this study was to evaluate whether crime changed at KIB-funded Project GreenSpaces sites relative to matched unfunded applicant sites. We used a matched-pair difference-in-differences event-study framework with crime data to compare changes at funded sites with contemporaneous changes at their matched comparison sites [18]. We focused primarily on Part I violent offenses during the first and second post-grant years, while evaluating total crime as a secondary outcome. In a complementary analysis, we used Dynamic Functional Coefficient Autoregressive Spatio-Temporal (DyFAST) modeling to characterize short-term temporal persistence within sites and spatial dependence across neighboring study sites [19].

This analysis was used to describe underlying violent-crime dynamics.

## 2. Methods

### 2.1 Study Cohort and Data Sources

We used a dataset centered on Indianapolis, Indiana, with crime observations spanning the period from 2011 to 2021 [20]. The intervention sites were Project GreenSpaces projects funded through Keep Indianapolis Beautiful (KIB), a nonprofit organization dedicated to aesthetic and ecological improvement [17]. Project GreenSpaces is a community-led urban greening initiative that transforms vacant or underutilized lots into functional community spaces. Project designs varied across sites and included combinations of vegetation and community-oriented site amenities, with local partners involved in ongoing maintenance and care [17]. Crime incident data were obtained from the Indianapolis Metropolitan Police Department (IMPD) through the Open Indy Data Portal [20]. Site coordinates for KIB-funded Project GreenSpaces locations and the year in which each project received funding were obtained from KIB project records.

Census-tract-level demographic and socioeconomic characteristics were obtained from the American Community Survey (ACS) using the tidycensus package in R and linked to study sites to characterize neighborhood context [21].

### 2.2 Study Design and Matching

To evaluate changes in crime associated with the greening intervention, we used a matched-pair design comparing funded Project GreenSpaces sites with matched comparison sites.

Comparison sites were unfunded applicant sites from KIB greening projects. From an initial candidate pool of 58 funded and 110 unfunded sites, comparison sites were selected using 1:1 propensity-score matching on baseline neighborhood sociodemographic characteristics, implemented using MatchIt function in R [22]. Propensity scores were based on measures of poverty, vehicle access, age composition, disability, limited English proficiency, racial composition, food-assistance eligibility, housing tenure and cost burden, and educational attainment. Two matched pairs involving school-district properties were subsequently excluded, leaving a parent cohort of 34 matched pairs (N = 68 sites) (**Figures 1 and 2**). For each pair, the grant year of the funded site served as the common index year to align the pre- and post-grant observation periods.

**Figure 1.**
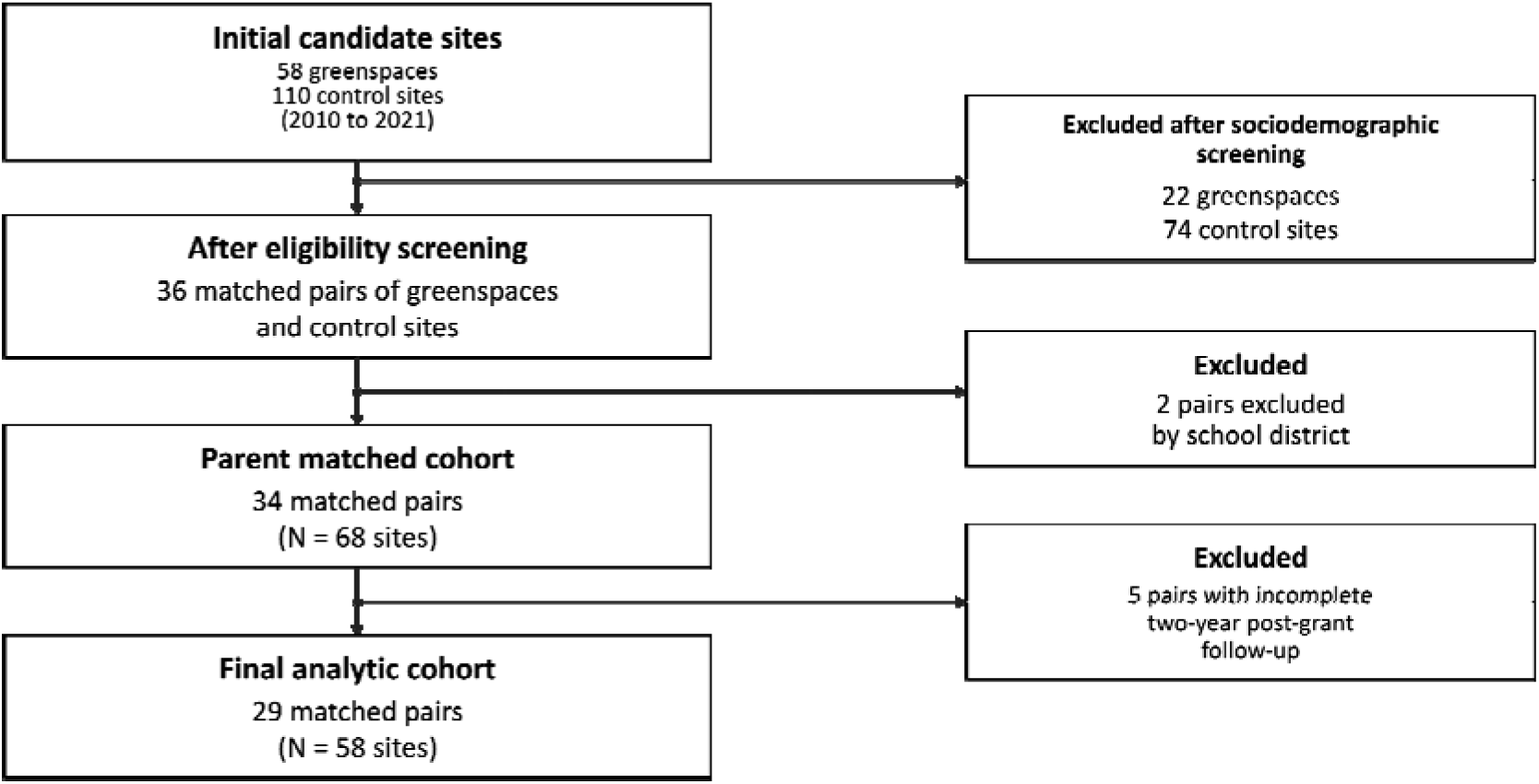
Flowchart of the selection and exclusion of sites. **Note:** Funded sites were KIB Project GreenSpaces sites, and comparison sites were unfunded applicant sites. Sites were matched 1:1 using propensity scores based on baseline neighborhood sociodemographic characteristics. The final analytic cohort included matched pairs with complete two-year pre- and post-grant follow-up.

**Figure 2.**
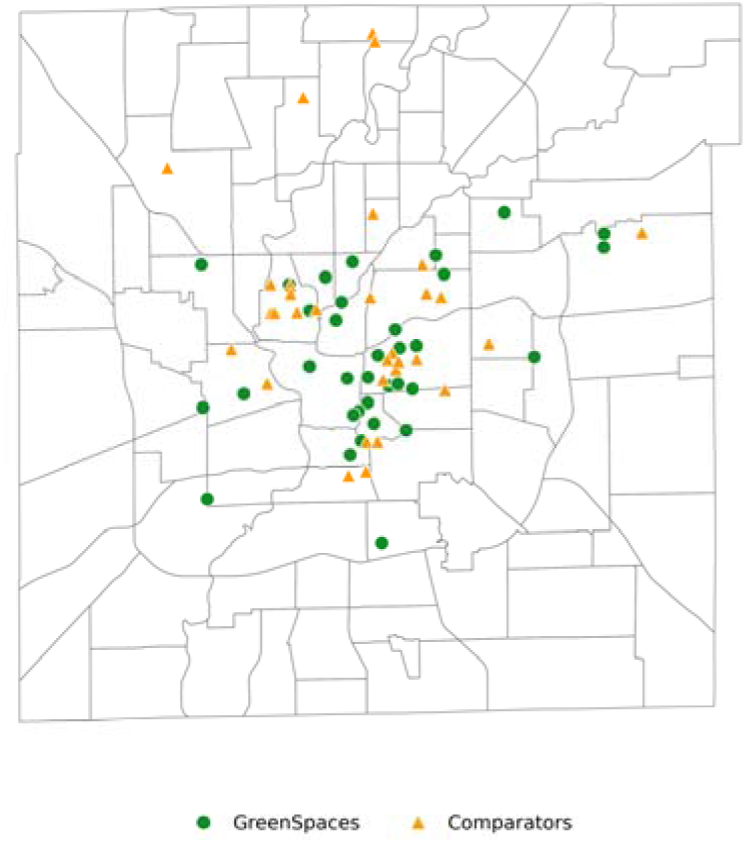
Distribution of greenspace sites and matched comparison sites. **Note:** Green circles represent funded Project GreenSpaces sites, and orange triangles represent matched unfunded comparison sites. The map includes all 34 matched pairs (68 sites).

For our primary analysis, we further restricted this parent cohort to 29 matched pairs (58 sites) with complete two-year pre- and post-grant observation periods. The remaining five pairs were excluded as their later grant years lacked the required two-year post-grant follow-up period. Baseline characteristics of the final 29-pair analytic sample are reported in **Table 1**.

**Table 1.** Baseline Sociodemographic Characteristics and Pre-grant Crime Outcomes Among 29 Matched Funded–Comparison Site Pairs.

| Characteristic (%) | GreenSpace<br>(n = 29)<sup>1</sup> | Comparators<br>(n = 29)<sup>1</sup> |
| --- | --- | --- |
| **Baseline sociodemographic characteristics** |  |  |
| Households below 200% poverty | 58.6 (18.5) | 64.6 (11.6) |
| No vehicle access | 20.9 (7.9) | 20.3 (7.4) |
| Population under 18 | 21.6 (9.0) | 24.8 (5.2) |
| Population 65 or older | 9.9 (4.8) | 9.7 (3.6) |
| Disability | 17.8 (5.0) | 18.6 (4.8) |
| Limited English proficiency | 2.9 (3.4) | 3.3 (3.1) |
| Non-White population | 43.0 (24.1) | 54.7 (27.3) |
| Food-stamp eligible households | 29.2 (12.4) | 31.2 (11.0) |
| Housing-cost burden | 32.9 (10.8) | 36.3 (11.2) |
| Renter-occupied housing | 59.6 (9.9) | 58.7 (11.6) |
| No high-school diploma | 25.0 (10.3) | 25.0 (9.4) |
| **Pre-grant outcome** |  |  |
| Part I violent offenses/month | 4.07 (3.50) | 4.95 (3.37) |
| Total crime/month | 16.86 (10.97) | 18.41 (10.91) |

<sup>1</sup> Mean (SD)
**Note:** Pre-grant crime measures represent each site's mean monthly count during the two years preceding the grant year, summarized at the site level.

### 2.3 Outcome Measures

Crime data were available at the incident level and geocoded to incident location. For each study site, crime incidents occurring within a 600-m circular buffer were identified and used to construct monthly site-level crime outcomes. Duplicate records of the same source incident within a site were removed; incidents falling within overlapping site buffers could contribute once to each affected site. The primary outcome was Part I violent offenses, including homicide, rape, robbery, and aggravated assault. The secondary outcome was total recorded crime, defined as all recorded crime incidents within the study-site buffer, without restriction to offense type.

### 2.4 Primary Analysis

To evaluate changes in crime associated with greening, we used a matched-pair difference-in-differences (DiD) event-study framework estimated via Poisson pseudo-maximum likelihood (PPML) [18]. The model compared monthly crime counts at funded sites with their matched comparison sites, controlling for time-invariant local characteristics (site fixed effects) and month-specific factors shared by the two sites within each matched pair (matched-pair-by-calendar year-month fixed effects). Event time was defined relative to the grant year of the funded site, which served as the common index year for both sites within each matched pair. Because the exact project completion month was unavailable, the grant year (year 0) was treated as a transition period and excluded from the event study. The preceding year (year −1) served as the reference period, and coefficients were estimated for year −2 and post-grant years +1 and +2. Estimates are reported as incidence rate ratios (IRRs) with 95% confidence intervals. Standard errors were clustered by matched pair to account for within-pair dependence, with inference additionally evaluated using a matched-pair bootstrap. Full model specification and inference procedures are provided in **eMethod 1**.

### 2.5. Sensitivity Analyses for the Primary Analysis

Robustness analyses evaluated residual baseline imbalance, potential spatial interference, follow-up completeness, model specification, and outcome radius. We fitted models including baseline sociodemographic covariate-by-relative-year interactions, repeated analyses using available observations from all 34 matched pairs and examined overlap-adjusted and overlap-free specifications. A narrower 300-m buffer was evaluated as a post hoc spatial-scale sensitivity analysis. Pre-grant temporal patterns were further examined using month-specific funded–comparison contrasts over the 24 months before the grant year, centered to the average contrast during months −12 to −1.

### 2.6. Secondary Analysis of Spatiotemporal Crime Dynamics

As a separate secondary analysis, we applied Dynamic Functional-Coefficient Autoregressive Spatio-Temporal (DyFAST) modeling to characterize the underlying spatial and temporal dynamics of crime outcomes [19]. We examined whether violent crime tended to persist from one month to the next at the same site (temporal persistence) and whether prior-month violent crime at nearby study sites was associated with subsequent crime at a given site (spatial dependence). DyFAST was used descriptively rather than to estimate the effect of greening and therefore did not include an intervention coefficient.

Monthly violent-crime counts within 600-m buffers were transformed as log(1 + count), and nearby sites were defined using an inverse-distance eight-nearest-neighbor structure. Primary results were summarized at the median of the broader crime-condition measure, with lower and higher values and alternative definitions of nearby sites examined in supplementary analyses. Full model specification and inference procedures are provided in **eMethod 2**.

## 3. Results

### 3.1 Baseline Sociodemographic Characteristics

The primary analytic sample comprised 29 matched funded–comparison site pairs with complete two-year pre- and post-grant follow-up. Baseline sociodemographic characteristics were similar across several measures, although residual differences remained in racial composition, age composition, poverty, and housing-cost burden **(Table 1)**. Pre-grant crime levels were more similar between groups. During the two years preceding the grant year, funded sites averaged 4.07 (SD = 3.50) Part I violent offenses per month, compared with 4.95 (SD = 3.37) at comparison sites; mean monthly total crime counts were similarly close at 16.86 (SD = 10.97) and 18.41 (SD = 10.91), respectively.

### 3.2. Primary Difference-in-Differences

During the first post-grant year, the violent-crime point estimate at greening sites was lower relative to the contemporaneous change at matched unfunded comparison sites, although the confidence interval included the null (IRR = 0.903, 95% CI 0.805–1.014, *P* = 0.083), corresponding to a 9.7% lower relative rate. In the second post-grant year, the estimated IRR was 0.830 (95% CI 0.718–0.961, *P* = 0.014), corresponding to a 17.0% lower relative rate **(Table 2).** The estimate for the second pre-grant year was close to the reference period (IRR = 0.985, 95% CI 0.892–1.087, *P* = 0.753).

**Table 2.** Difference-in-Differences Estimates for Crime Outcomes.

| **Outcome** | **Relative year** | **IRR (95% CI)** | **% change** | **p-value** |
| --- | --- | --- | --- | --- |
| **Part I violent offenses** | Year -2 | 0.985 (0.892–1.087) | –1.5% | 0.753 |
|  | Post-grant year 1 | 0.903 (0.805–1.014) | –9.7% | 0.083 |
|  | Post-grant year 2 | 0.830 (0.718–0.961) | –17.0% | 0.014 |
| **Total crime** | Year -2 | 0.999 (0.921–1.083) | –0.1% | 0.974 |
|  | Post-grant year 1 | 0.974 (0.890–1.065) | –2.6% | 0.546 |
|  | Post-grant year 2 | 0.986 (0.879–1.107) | –1.4% | 0.807 |

**Note:** Estimates are from Poisson pseudo-maximum likelihood difference-in-differences models with site fixed effects and matched-pair-by-calendar year-month fixed effects. Year –1 served as the reference period, and the grant year (year 0) was excluded as a transition period. IRRs compare relative changes at funded sites with contemporaneous changes at matched comparison sites.
IRR = incidence rate ratio; CI = confidence interval; PPML = Poisson pseudo-maximum likelihood.

In contrast, total recorded crime did not show a clear post-grant decrease. Estimates remained close to the null in the first post-grant year (IRR = 0.974, 95% CI 0.890–1.065, *P* = 0.546) and the second post-grant year (IRR = 0.986, 95% CI 0.879–1.107, *P* = 0.807). The corresponding year −2 estimate was also close to the null (IRR = 0.999, 95% CI 0.921–1.083, *P* = 0.974).

### 3.3 Sensitivity Analyses

Across alternative specifications, sensitivity analyses were more consistent for the second-year violent crime reduction **(ST1)**. Adjusting for baseline sociodemographic attenuated the first-year estimate (IRR, 0.935; 95% CI, 0.843–1.036), whereas the second-year estimate remained significant (IRR, 0.833; 95% CI, 0.733–0.946). Analyses of all 34 original pairs and overlap-adjusted and overlap-free specifications produced generally similar directional patterns. Additional model and inference checks were also consistent with the primary direction.

Spatial-scale analyses indicated that estimates were sensitive to the outcome radius. With 300-m buffers, the first-year estimate was close to the null and slightly above one (IRR, 1.061; 95% CI, 0.771–1.461), whereas the second-year estimate remained below one but was substantially less precise (IRR, 0.794; 95% CI, 0.569–1.107). Total crime did not show a clear decrease at the 300-m scale.

Monthly pre-grant diagnostics showed that funded–comparison contrasts fluctuated around the pre-grant average without a sustained directional pattern at 600 m (**SF1**). Estimates at 300 m were less precise and more variable, with no persistent pre-grant pattern.

### 3.4 Temporal Persistence and Spatial Dependence of Violent Crime

The DyFAST analysis focused on Part I violent crime, for which the matched DiD analysis showed the post-grant association, to further characterize its short-term temporal and spatial dynamics (Table 3). Under the primary specification, the site-averaged temporal coefficient was 0.418 (95% CI, 0.383–0.450), indicating that higher violent-crime levels in the preceding month were associated with higher levels in the subsequent month at the same site (**Table 3**). The corresponding neighboring-site spatial coefficient was 0.052 (95% CI, −0.002–0.113), providing limited evidence of a stable one-month spatial association under the primary specification. Overall, the DyFAST analysis demonstrated that short-term violent-crime patterns exhibited more consistent within-site temporal persistence than associations between prior-month violent crime at nearby sites and subsequent violent crime at a given site.

**Table 3.**
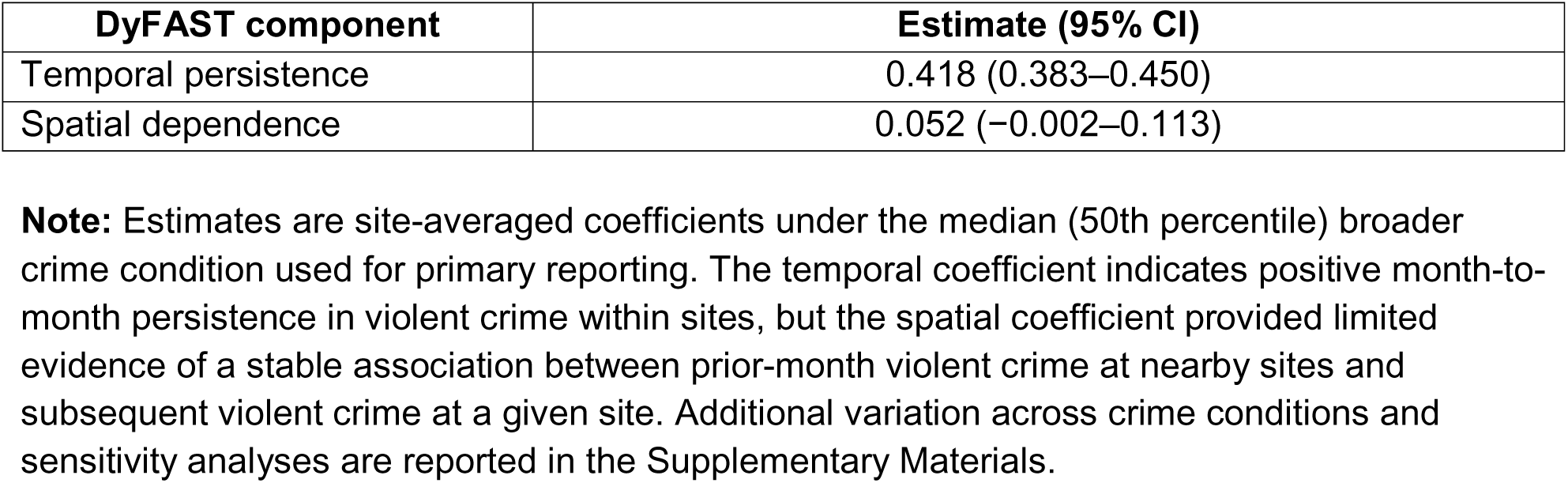
Secondary DyFAST Estimates of One-Month Violent-Crime Dynamics.

## 4. Discussion

In this matched study of community-led greening projects in Indianapolis, the primary 600-m analysis showed a modest, imprecisely estimated decrease in Part I violent crime during the first post-grant year and a clearer relative decrease during the second. The estimated relative rate was approximately 10% lower in the first post-grant year and 17% lower in the second, with the second-year association more consistent across sensitivity analyses. In contrast, we found little evidence of a corresponding change in total crime. Thus, these findings suggest that, within the two-year post-grant follow-up, the association between Project GreenSpaces and subsequent violent-crime patterns was most evident in the second post-grant year.

The greater consistency of the second-year estimate is broadly consistent with prior longitudinal evidence suggesting that crime-related changes following resident-led greening may evolve over time. Hadavi et al. reported reductions in crime beginning in the second year of implementation of a resident-led vacant-lot reuse program, while a community-engaged greening study by Gong et al. also reported declines in violent crime [23, 24]. However, the present study followed sites for only two post-grant years and therefore cannot determine whether the observed pattern would persist, strengthen, attenuate, or otherwise change with longer follow-up. The absence of exact implementation and completion dates also introduces uncertainty regarding the timing of changes relative to physical site transformation.

The sensitivity analyses revealed some variation across spatial and temporal scales. At 600 m, the second-post-grant-year estimate remained relatively stable across alternative specifications, whereas the first-year estimate was more sensitive. When the outcome radius was reduced to 300 m, the first-year association was attenuated, and the second-year estimate became less precise. Monthly pre-grant diagnostics showed no sustained directional pattern at 600 m; estimates at 300 m were more variable but likewise did not indicate a persistent pre-grant pattern. The contrast across these analyses indicates that the observed associations depend partly on how local crime is defined in space and time.

The contrasting findings for violent and total crime provide additional context for interpreting the results. Lower relative rates after the grant year were observed for Part I violent offenses, whereas total-crime estimates remained close to the null. The decrease observed for Part I violent offenses was not mirrored in the aggregated total crime outcome. Total crime combines offenses with different behavioral and spatial patterns, and changes in a particular category may therefore be less apparent when all offenses are aggregated. The findings are therefore more consistent with a selective change in violent-crime patterns than with a generalized change in crime around the study sites.

The DyFAST analysis provided complementary context alongside the matched DiD analysis by characterizing short-term violent-crime dynamics across the study sites. Violent crime showed clear month-to-month persistence within sites, whereas associations between prior-month violent crime at nearby sites and subsequent violent crime at a given site were smaller and sensitive to the spatial-network definition. Because DyFAST did not estimate an intervention effect, these findings should not be interpreted as confirming the DiD estimates or identifying a mechanism linking Project GreenSpaces to post-grant crime patterns. Its role here is to provide additional context for the temporal and spatial structure of violent crime.

Several features strengthen this study, including the use of unfunded applicants as comparison sites, repeated monthly crime observations, and a matched longitudinal design that allowed funded and comparison sites to be followed around a common grant year. The analysis also examined whether the results changed under alternative spatial scales, follow-up definitions, and specifications. These strengths notwithstanding, several important limitations warrant consideration. Funding was not randomly assigned, some baseline differences persisted after matching, and only the grant year—not the dates of project implementation or completion—was available. The two-year post-grant follow-up also captures a relatively early period in the trajectory of these projects. Project GreenSpaces has continued beyond the observation window examined here, and longer-term associations may differ as sites mature, maintenance practices change, and patterns of community use evolve. Police-recorded incidents may reflect reporting and enforcement practices as well as underlying crime and overlapping site buffers complicate the assumption that each location was completely independent of nearby projects.

In conclusion, this matched study observed lower relative violent-crime rates during the two-year post-grant period, with the most consistent association in the second year. These findings characterize short-term post-grant patterns rather than long-term program effects of this community-led greening initiative in Indianapolis. Longer follow-up will be important to determine whether these associations persist or change as projects mature. Future work would benefit from exact implementation dates and direct measures of site maintenance, use, and resident engagement, which could help distinguish whether changes in crime accompany the physical transformation of a site, its subsequent use by residents, or both.

## Supporting information

supp.docx

## Data Availability

Indianapolis crime data were obtained from the Open Indy Data Portal. Census-tract-level sociodemographic variables were obtained from the U.S. Census Bureau’s American Community Survey (ACS) using the tidycensus R package.

## Acknowledgments

We gratefully acknowledge Keep Indianapolis Beautiful (KIB) for providing the greenspace project data and for its support of this work. We thank Karl Selm, Kelly Cook, Katherine Waters, and Carly Weidman of KIB for their contributions to the project. We also acknowledge the City of Indianapolis and AES Indiana, the title sponsor of KIB’s greenspace program, for their support of the program. Finally, we thank Kathleen Howe, Elizabeth Gonzalez, and Kai Le of the University of Michigan for their contributions to this work.

## Supplementary Information

Supplementary Information is available for this paper.

## Abbreviations

ACS: American Community Survey;
CI: confidence interval;
DiD: difference-in-differences;
DyFAST: Dynamic Functional-Coefficient Autoregressive Spatio-Temporal;
IMPD: Indianapolis Metropolitan Police Department;
IRR: incidence rate ratio;
KIB: Keep Indianapolis Beautiful;
PPML: Poisson pseudo-maximum likelihood;
SD: standard deviation

