## Supplementary material for "Spatial and Temporal Evaluation of Violent and Total Crime Following Community-Led Urban Greening in Indianapolis": supp.docx

**eMethod1. Primary Difference-in-Differences Specification**

Let $Y_{ipt}$ denote the monthly crime count for site $i$in matched pair $p$ during the calendar month $t$, and let $G_{i}$ indicate a funded site. Relative event year was defined as $r= calendar year-grant year$. The grant year ($r=0$) was omitted as a transition period, and $r=-1$served as the reference period.

The primary PPML model was:

$$logE\left( Y_{ipt} \right)=\alpha_{i}+\eta_{pt}+\beta_{-2}G_{iI\left( r=-2 \right)}+\beta_{1}G_{iI\left( r=+1 \right)}+\beta_{2}G_{iI\left( r=+2 \right)}$$

where $\alpha_{i}$ denotes site fixed effects, and $\eta_{pt}$ denotes matched-pair-by-calendar year-month fixed effects. Thus, identification derives from differential changes between the two members of a matched pair within the same calendar year-month. Exponentiated event-time coefficients were interpreted as incidence rate ratios (IRRs), representing the relative change at funded sites compared with the contemporaneous change at their matched comparison sites, using year −1 as the reference period.

Standard errors were clustered at the matched-pair level. Primary confidence intervals and *P* values used small-sample *t*-based inference. As an additional inference check, a nonparametric matched-pair cluster bootstrap was performed using 1,999 resamples, with both sites in a matched pair resampled together. Percentile and basic bootstrap confidence intervals were calculated separately and were used as sensitivity analyses rather than replacing the primary pair-clustered confidence intervals.

In sensitivity analyses, baseline sociodemographic covariates were interacted with event-time indicators because their time-invariant main effects were absorbed by the site fixed effects. We also examined analyses using all 34 matched pairs with available follow-up, and overlap-adjusted and overlap-free specifications to assess potential spatial interference. A narrower 300-m outcome definition was evaluated as a spatial-scale sensitivity analysis. Furthermore, month-specific funded–comparison contrasts were estimated during the 24 months preceding the grant year. To avoid selecting a single month as the reference, these contrasts were centered to the mean funded–comparison contrast during months −12 to −1, corresponding to the year −1 reference period in the annual DiD analysis.

**eMethod2. Analysis for Temporal Persistence and Spatial Dependence**

DyFAST was used as a secondary descriptive analysis to characterize short-term spatial and temporal patterns in monthly Part I violent crime rather than to estimate the effect of greening. 600-m monthly violent-crime counts were transformed as log(1 + count). Temporal persistence was represented by each site's violent-crime level in the preceding month, whereas spatial dependence was represented by an inverse-distance-weighted average of prior-month violent crime among the eight nearest study sites.

DyFAST allows these temporal and spatial relationships to vary across study locations and over broader crime conditions. The broader crime-condition measure was constructed from prior-month total-crime patterns across the other observed study sites and was distinct from the nearby-site violent-crime measure used to estimate spatial dependence. Specifically, log-transformed total crime was centered around each site's own average before being summarized across the other study sites and standardized.

To summarize how the estimated relationships varied with broader crime conditions, coefficients were evaluated at the 25th, 50th, and 75th percentiles of this measure, referred to as lower, median, and higher crime conditions, respectively. The main-text results report the median condition, while the lower and higher conditions are provided in the Supplementary Table to describe variation in the estimates. The labels lower, median, and higher therefore refer to relative levels of the constructed crime-condition measure rather than to categories of neighborhoods or absolute crime counts.

Bandwidths were selected using five-fold cross-validation, with the two sites in each matched pair assigned to the same fold. Models additionally accounted for baseline site characteristics, calendar-time trend, and annual seasonality. Confidence intervals were estimated using a linearized wild-multiplier procedure with calendar-time blocks. Final inference was based on 4,999 multiplier draws. Six-month blocks were used for primary reporting. The primary specification used eight neighboring sites; analyses using five and 10 neighbors and an alternative calendar-time block length were examined as sensitivity analyses.

**ST1. Sensitivity Analyses for Part I Violent and Total Crime**

| **Type** | **Analysis** | **Analytic sample** | **Post year 1 IRR (95% CI)** | **Post year 2 IRR (95% CI)** |
| --- | --- | --- | --- | --- |
| **Part I violent crime** | Primary analysis, 600 m | 29 pairs | **0.903 (0.805–1.014)** | **0.830 (0.718–0.961)** |
|  | Baseline covariate adjustment | 29 pairs | 0.935 (0.843–1.036) | 0.833 (0.733–0.946) |
|  | Matched-pair bootstrap inference | 29 pairs | 0.903 (0.808–1.010) | 0.830 (0.716–0.940) |
|  | Overlap-exposure adjustment | 29 pairs | 0.915 (0.835–1.004) | 0.845 (0.752–0.950) |
|  | Overlap-free subset | 16 pairs | 0.930 (0.797–1.084) | 0.850 (0.664–1.087) |
|  | All original pairs with available follow-up | 34 pairs | 0.874 (0.775–0.985) | 0.814 (0.704–0.942) |
|  | 300-m outcome definition | 29 pairs | 1.061 (0.771–1.461) | 0.794 (0.569–1.107) |
| Total crime | Primary analysis, 600 m | 29 pairs | 0.974 (0.890–1.065) | 0.986 (0.879–1.107) |
|  | 300-m outcome definition | 29 pairs | 1.095 (0.982–1.220) | 1.077 (0.885–1.310) |

**Note.** IRR = incidence rate ratio; CI = confidence interval; PPML = Poisson pseudo-maximum likelihood.

**ST2. DyFAST estimates and sensitivity analyses**

**Panel A. Variation across broader crime conditions**

| **Broader crime condition** | **Temporal coefficient (95% CI)** | **Spatial coefficient (95% CI)** |
| --- | --- | --- |
| Lower (25th percentile) | 0.408 (0.374–0.439) | 0.058 (0.011–0.109) |
| Median (50th percentile) | 0.418 (0.383–0.450) | 0.052 (−0.002–0.113) |
| Higher (75th percentile) | 0.425 (0.385–0.462) | 0.048 (−0.020–0.119) |

**Note.** These categories describe relative variation in crime conditions across the study sites and do not represent classifications of neighborhoods or absolute crime levels. Main-text results report estimates at the median condition.

**Panel B. Sensitivity to the number of neighboring sites**

|  | **Temporal coefficient (95% CI)** | **Spatial coefficient (95% CI)** |
| --- | --- | --- |
| 5 nearest sites | 0.404 (0.335–0.467) | 0.031 (−0.036–0.107) |
| 8 nearest sites (primary) | 0.418 (0.383–0.450) | 0.052 (−0.002–0.113) |
| 10 nearest sites | 0.429 (0.398–0.459) | 0.079 (0.024–0.143) |

**Note.** Results were similar when an alternative calendar-time block length was used for inference.

**SF1.** **Monthly Pre-Grant Funded–Comparison Contrasts in Part I Violent Crime**

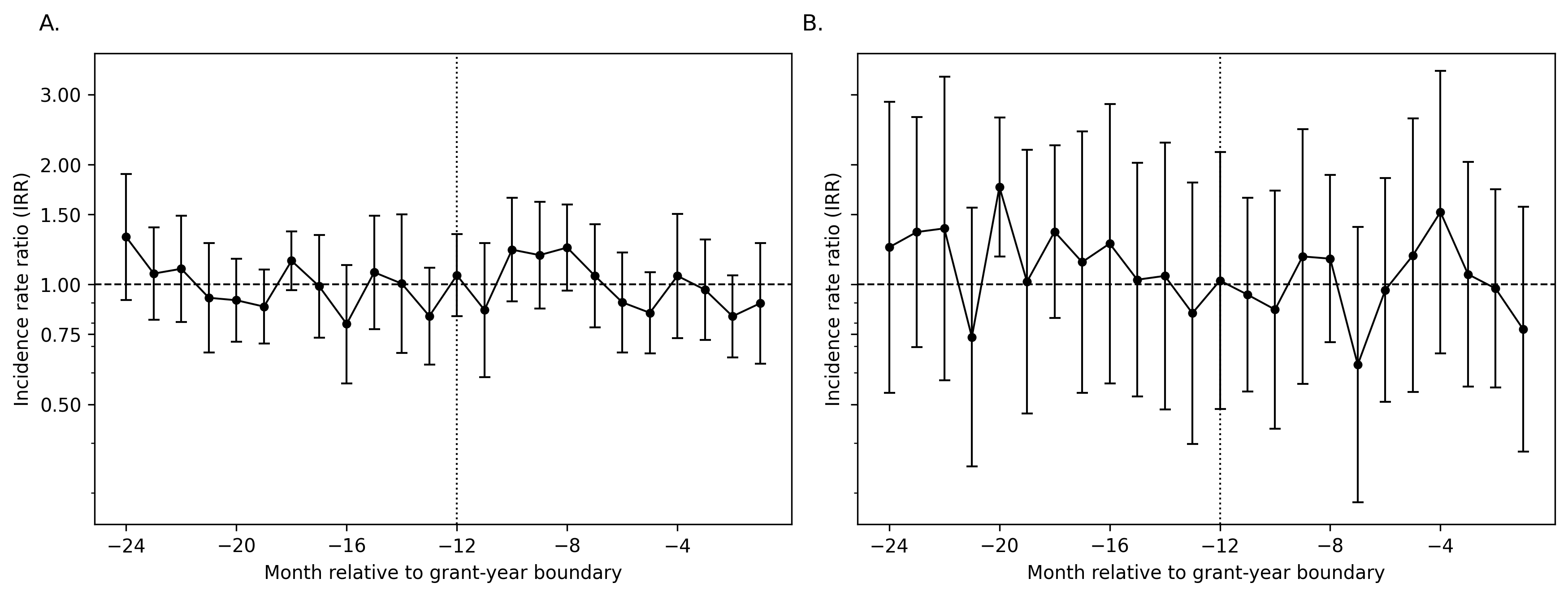

**Note**: Points indicate month-specific incidence rate ratios (IRRs) with 95% confidence intervals. Months are indexed relative to the start of the grant year. IRRs were centered to the average funded–comparison contrast during months −12 to −1, consistent with the year −1 reference period in the primary DiD analysis. IRR = 1 represents that average pre-grant contrast. Panel A, 600 m; Panel B, 300 m.

IRR = incidence rate ratio; DiD = Difference-in-Differences
